# Clinical selectivity and failure modes of automated chest radiograph report evaluation metrics: a cross-dataset analysis of ReXErr-v1 and RadEvalX

**DOI:** 10.64898/2026.08.10.26360043

**Authors:** Jatin Sridhar Naidu, Sandeep Muralidharan, Anuja Prashani, Vasanth Baskaradoss

## Abstract

**Objectives:** To test whether radiology report evaluation metrics distinguish clinically meaningful errors from textual changes and align with radiologist-assessed error burden.

**Methods:** Cross-dataset evaluation used ReXErr-v1 (2,708 report pairs; 5,724 paired error sentences) and 100 RadEvalX report pairs with expert error counts. BLEU-4, ROUGE-L and METEOR were assessed in ReXErr-v1; RadEvalX analyses included these plus BERTScore, CheXbert, RadGraph F1 and RadCliQ. Outcomes were ReXErr-v1 pairwise win rate and AUROC for clinical-content versus linguistic errors, and RadEvalX Spearman correlation with clinically significant error count and AUROC for any significant error. Confidence intervals used 10,000 clustered percentile bootstrap resamples; Holm adjustment-controlled multiplicity.

**Results:** ReXErr-v1 paired-sentence win rates were 0.986 for BLEU-4, 0.999 for ROUGE-L and 0.998 for METEOR, but discrimination of clinical-content from linguistic errors was modest (AUROC 0.609–0.620). Penalty magnitude was strongly associated with textual change after adjustment for error type (normalised character edit distance coefficient 0.746; 95% CI 0.705–0.788; P<0.001). In RadEvalX, CheXbert showed the highest correlation with clinically significant errors (rho=0.413; 95% CI 0.223–0.578) and highest AUROC (0.742; 95% CI 0.638–0.836).

**Conclusions:** Near-ceiling sensitivity to textual corruption did not imply sensitivity to clinical significance. CheXbert showed the highest alignment with expert error assessment, although pairwise superiority was not demonstrated over all comparators and performance remained moderate.

**Advances in knowledge:** This study separates textual corruption sensitivity from clinical selectivity across complementary synthetic and expert-annotated benchmarks, showing that apparent error-detection performance can be strongly influenced by textual change rather than clinical importance alone.

## Introduction

Automated chest radiograph report generation is increasingly evaluated using quantitative measures of agreement between generated and reference reports. Conventional metrics such as BLEU,^1^ ROUGE,^2^ and METEOR primarily reward lexical overlap,^3^ whereas embedding- and radiology-aware approaches including BERTScore, CheXbert, RadGraph-based scores and RadCliQ aim to capture progressively more semantic or clinically relevant information.^4-7^ More recent radiology-specific approaches, including RaTEScore and GREEN, explicitly target medical entities or clinically important report errors.^8,9^

Currently, RadEval provides a unified framework spanning lexical, semantic, clinically structured and large language model-based metrics, including BLEU, ROUGE, BERTScore, CheXbert, RadGraph, RaTEScore and GREEN, together with expert-labelled evaluation.^10^ ReFINE introduced an interpretable reward-based framework designed to improve alignment with human assessment,^11^ while GEMA-Score uses granular multi-agent evaluation and has demonstrated alignment with expert judgements on datasets including RadEvalX.^12^ Most recently, RadOT-Eval evaluated a structured-evidence transport metric using expert annotations from RadEvalX and a corruption-sensitivity stress test on ReXErr-v1.16.^13^ Collectively, these studies have substantially advanced metric benchmarking and expert-alignment assessment.

However, alteration sensitivity is not equivalent to clinical selectivity. A metric may reliably detect that a report has changed without preferentially penalising changes that matter clinically. For example, it may penalise a typographical or homophone alteration as readily as a false negation, or its penalty may primarily reflect the amount of edited text rather than the clinical consequence of the alteration. Conversely, a clinically oriented metric may be relatively insensitive to superficial wording changes while better reflecting radiologist judgement. This distinction is important when automated metrics are used to compare report-generation models, select checkpoints or support claims of clinical fidelity.^14^

Accordingly, the contribution of the present study is not another leaderboard of competing report-evaluation metrics. Rather, we examine how and why metric scores change by separating the ability to detect an alteration from the ability to prioritise clinically meaningful alterations. Specifically, we evaluate clinical-versus-linguistic perturbation selectivity, identify error categories that produce no metric penalty, and test whether penalty magnitude is associated with the magnitude of the underlying textual edit. These analyses complement recent expert-correlation and benchmark studies by interrogating metric failure modes that aggregate correlations or discrimination statistics alone may obscure.

Two public chest radiograph report datasets provide complementary settings for this analysis.

ReXErr-v1 contains synthetic error-injected reports derived from MIMIC-CXR, with report-level and sentence-level error information.^15,16^ RadEvalX contains 100 model-generated reports compared with IU-Xray reference reports and consensus annotations from two board-certified radiologists who counted clinically significant and clinically insignificant errors.^17,18^ Although both datasets have recently been used in metric-development and benchmarking studies,^13^ their complementary structures permit the present analysis to distinguish controlled alteration sensitivity from alignment with expert-assessed clinical error burden.

We therefore evaluated the clinical selectivity and failure modes of automated report metrics across ReXErr-v1 and RadEvalX. The objectives were to: (1) quantify how consistently lexical metrics detect report- and sentence-level corruption in ReXErr-v1; (2) determine whether penalty magnitude distinguishes clinical-content from linguistic errors, characterise category-specific no-penalty failures, and assess whether penalties are associated with edit magnitude; and (3) compare available metrics against expert clinically significant error burden in RadEvalX. We hypothesised that near-perfect detection of synthetic textual alterations would not necessarily translate into strong clinical selectivity or expert alignment.

## Methods

### Study design and data sources

This was a secondary methodological evaluation of two previously released, de-identified chest radiograph report datasets. ReXErr-v1 version 1.0.0 is derived from MIMIC-CXR and contains original reports paired with reports containing synthetic errors, together with sentence-level post-hoc error labels.^15,16^ The analysed report-level test set comprised 2,708 original/error-injected report pairs. The sentence-level test file contained 19,859 rows: 10,790 labelled no error, 8,723 labelled error and 346 neutral. Every report-level test case contained exactly three sampled error categories, so report-level performance could not be stratified by the number of injected errors.

RadEvalX version 1.0.0 comprises 100 M2Tr-generated report/reference-report pairs sampled from IU-Xray.^17,18^ Two board-certified radiologists independently assessed the pairs and reached consensus on counts of clinically significant and clinically insignificant errors across eight categories. The supplied metric file contained BLEU-4, BERTScore, CheXbert similarity, RadGraph F1 and RadCliQ values. Sixty-two of the 100 reports contained at least one clinically significant error.

### Analytical populations and error grouping

For ReXErr-v1 report-level analyses, the original report was treated as the reference and both the original and error-injected report were scored against it. Sentence-level paired analyses were restricted to error-labelled rows with both an original and altered sentence. Of 8,723 error-labelled rows, 2,523 lacked an original sentence and 476 lacked an altered sentence, leaving 5,724 complete pairs. Unpaired insertion and deletion cases were excluded rather than represented by a literal missing-value token (**Table 1**).

**Table 1.** Data sources and analytical populations. ReXErr-v1 sentence-level paired rows include both error-labelled and no-error rows where both text fields were present. The primary paired error analysis was restricted to error-labelled complete pairs.

| Dataset/component | Available rows | Analysed rows | Key feature |
| --- | --- | --- | --- |
| ReXErr-v1 report-level test | 2,708 | 2,708 | Each report contained exactly three sampled error categories |
| ReXErr-v1 sentence-level test | 19,859 | 16,860 paired rows | 10,790 no-error; 8,723 error; 346 neutral |
| ReXErr-v1 paired error analysis | 8,723 error-labelled | 5,724 complete pairs | 2,999 rows lacked original or altered sentence |
| RadEvalX | 100 | 100 | 62 reports had $\geq 1$ clinically significant error |

For the clinical-selectivity analysis, paired error sentences were grouped by error type. Clinical-content errors comprised change location, change measurement, change device name, change device position, change severity, false negation and false prediction (n=4,463). Linguistic errors comprised added typographical errors and homophone substitutions (n=1,258). The ReXErr-v1 clinical-content grouping reflects the semantic nature of the injected error rather than expert-rated clinical significance;^15,16^ clinical significance was assessed only in RadEvalX.^17^ Three complete add-medical-device pairs were retained in overall and category-specific analyses but excluded from the grouped clinical-versus-linguistic comparison because of sparse representation. Rows labelled as containing no error were retained as a negative-control audit of sentence alignment and post-hoc labelling.

### Evaluation metrics

BLEU-4, ROUGE-L and METEOR were calculated locally for ReXErr-v1 using the implementations described below.^1-3^ For RadEvalX, ROUGE-L and METEOR were likewise calculated locally from the candidate/reference report pairs, while the supplied BLEU-4 values were used after confirming that the local BLEU-4 implementation reproduced them to a maximum absolute difference below 5 × 10™10. BLEU-4 used lower-cased whitespace tokenisation, equal four-gram weights and no smoothing.^1^ ROUGE-L used the longest-common-subsequence F1 score.^2^ METEOR used exact and Porter-stem matches without WordNet synonym matching because the required WordNet resource was unavailable; results therefore refer specifically to this exact-and-stem implementation.^3^

RadEvalX additionally provided BERTScore, CheXbert similarity, RadGraph F1 and RadCliQ values, which were analysed as supplied and were not independently recomputed.^17^ Higher BLEU-4, ROUGE-L, METEOR, BERTScore, CheXbert and RadGraph F1 indicated greater similarity or report quality, whereas higher RadCliQ indicated greater error burden. For expert-alignment analyses, metric direction was harmonised so that larger oriented values represented greater error burden.

### Outcomes and statistical analysis

The primary ReXErr-v1 outcome was pairwise win rate. For metric penalty Δ=Qoriginal™Qerror, a win was scored as 1 when Δ>1×10™12, 0.5 for a numerical tie (|Δ|≤1×10™12) and 0 when Δ<™1×10™12; metric scores were not rounded before comparison. Because the original report or sentence was evaluated against itself as the reference condition, this outcome primarily measures sensitivity to introduced textual perturbation rather than clinical error detection. Secondary outcomes were no-penalty rate and penalty magnitude. Confidence intervals were estimated from 10,000 non-parametric percentile bootstrap replicates using parent report as the ReXErr-v1 resampling unit, thereby preserving within-report clustering. A fixed random seed of 20260806 was used.

Clinical selectivity was assessed by the AUROC for discriminating clinical-content from linguistic errors using penalty magnitude. Clustered sentence-level modelling used a Gaussian generalised estimating equation (GEE) with an exchangeable working correlation structure and parent report as the clustering unit.^19^ Penalty was transformed separately within each metric to its average percentile rank. Fixed effects included metric, error type, their interaction, original sentence length, absolute token change and normalised character edit distance. Tokens were lower-cased whitespace-delimited units; absolute token change was the absolute difference in token counts between erroneous and original sentences. Normalised character edit distance was the lower-cased character-level Levenshtein distance divided by the larger of the two character lengths (minimum denominator 1). An initial logistic GEE of binary error detection did not converge because ROUGE-L and METEOR showed near-complete detection, producing quasi-complete separation.

The primary RadEvalX outcome was Spearman correlation between each oriented metric error signal and the count of clinically significant errors. Secondary outcomes were AUROC and average precision (AP) for at least one clinically significant error, correlation with clinically insignificant error count, and the difference between significant- and insignificant-error correlations. RadEvalX confidence intervals used 10,000 report-level percentile bootstrap samples. Differences between correlations and paired metric comparisons were bootstrapped using the same resampled reports for both statistics; two-sided empirical bootstrap P values were Holm-adjusted within defined comparison families. Analyses were performed in Python 3.13.5 using NumPy 2.3.5, pandas 2.2.3, SciPy 1.17.0, scikit-learn 1.8.0, statsmodels 0.14.6, NLTK 3.9.2 and RapidFuzz 3.14.3.^20-23^ No statistical imputation was performed. Paired ReXErr-v1 analyses required both text fields; blank RadEvalX error-category cells were treated as zero counts, and metric comparisons used observations available for both metrics. Full computational definitions, package functions and missing-data rules are provided in the **Supplementary Material 1. Supplementary Material 2** provides detailed analysis results, dataset flow, metric performance tables, category-specific analyses, pairwise metric comparisons, GEE model outputs, negative-control audits and quality-control/deviation logs.

The analyses were not prospectively preregistered and are therefore considered exploratory secondary analyses.

### Ethics

This study involved secondary analysis of previously released de-identified datasets and no new participant recruitment or collection of identifiable patient data. ReXErr-v1 derives from MIMIC-CXR; the dataset custodians report institutional review board approval for the source MIMIC datasets at the Massachusetts Institute of Technology (protocol 0403000206) and Beth Israel Deaconess Medical Center (protocol 2001-P-001699/14).^15^ RadEvalX is based on de-identified IU-Xray reports.^17^

## Results

### Dataset audit

The 5,724 evaluable paired ReXErr-v1 error sentences represented 65.6% of the 8,723 error-labelled rows. Missing original sentences were concentrated in insertion-type errors, while 476 missing altered sentences occurred in false-negation rows. Among 10,790 rows labelled as containing no error, 419 (3.88%) nevertheless contained non-identical original and altered text. BLEU-4 and METEOR penalised 2.62% of these nominal no-error rows and ROUGE-L penalised 2.69%, indicating residual sentence alignment or post-hoc label noise.

### Sensitivity to textual perturbation in ReXErr-v1

At report level, BLEU-4 produced a pairwise win rate of 0.9994 (95% CI 0.9987-1.0000), while ROUGE-L and METEOR each achieved 1.0000. BLEU-4 produced three numerical ties; ROUGE-L and METEOR penalised every error-injected report. Median penalties were 0.2350 for BLEU-4, 0.1179 for ROUGE-L and 0.0813 for METEOR (**Table 2; Figure 1**).

**Table 2.** ReXErr-v1 report- and paired-sentence perturbation-detection performance.

| Level | Metric | n | Pairwise win rate (95% CI) | No-penalty rate | Median penalty (IQR) |
| --- | --- | --- | --- | --- | --- |
| Report | BLEU-4 | 2,708 | 0.9994 (0.9987-1.0000) | 0.0011 | 0.2350 (0.1725-0.3162) |
| Report | ROUGE-L | 2,708 | 1.0000 (1.0000-1.0000) | 0.0000 | 0.1179 (0.0811-0.1715) |
| Report | METEOR | 2,708 | 1.0000 (1.0000-1.0000) | 0.0000 | 0.0813 (0.0500-0.1325) |
| Sentence | BLEU-4 | 5,724 | 0.9863 (0.9841-0.9884) | 0.0274 | 0.2983 (0.1718-0.6072) |
| Sentence | ROUGE-L | 5,724 | 0.9991 (0.9985-0.9997) | 0.0017 | 0.1304 (0.0769-0.2500) |
| Sentence | METEOR | 5,724 | 0.9979 (0.9970-0.9987) | 0.0042 | 0.1342 (0.0770-0.2561) |

**Figure 1.**
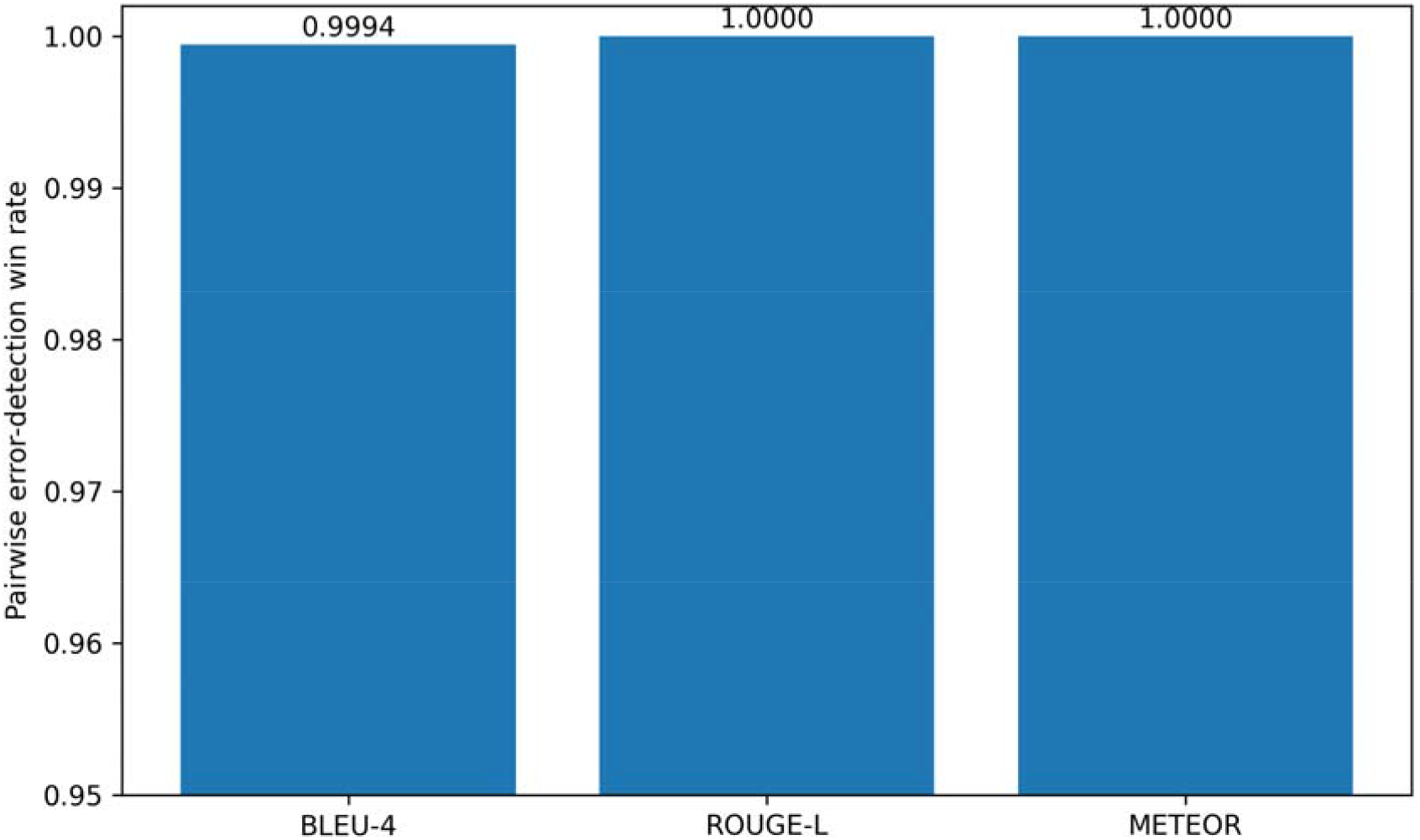
ReXErr-v1 report-level pairwise perturbation-detection win rates. BLEU-4, ROUGE-L and METEOR were used to compare each error-injected report with its original reference report. Win rates were 0.9994 for BLEU-4 and 1.0000 for both ROUGE-L and METEOR, indicating near-ceiling sensitivity to introduced textual perturbations at report level. This outcome reflects perturbation detection rather than clinical significance. **Alt text:** Bar chart showing ReXErr-v1 report-level pairwise perturbation-detection win rates for BLEU-4, ROUGE-L and METEOR; rates were 0.9994, 1.0000 and 1.0000, respectively, indicating near-ceiling detection across all three metrics.

At sentence level, win rates were 0.9863 (95% CI 0.9841-0.9884) for BLEU-4, 0.9991 (0.9985-0.9997) for ROUGE-L and 0.9979 (0.9970-0.9987) for METEOR. No erroneous sentence scored better than its original counterpart for any of the three metrics. BLEU-4 assigned no penalty to 2.74% of paired error sentences, with the highest no-penalty rates for change severity (4.48%), false negation (4.45%) and false prediction (4.25%).

### Clinical-content versus linguistic selectivity

Clinical-content errors received larger median penalties than linguistic errors, but discrimination was modest. AUROC was 0.612 (95% CI 0.595-0.629) for BLEU-4, 0.620 (0.605-0.634) for ROUGE-L and 0.609 (0.593-0.624) for METEOR (**Table 3**). Median clinical versus linguistic penalties were 0.331 versus 0.239 for BLEU-4, 0.143 versus 0.111 for ROUGE-L and 0.145 versus 0.104 for METEOR (**Figure 2**).

**Table 3.**
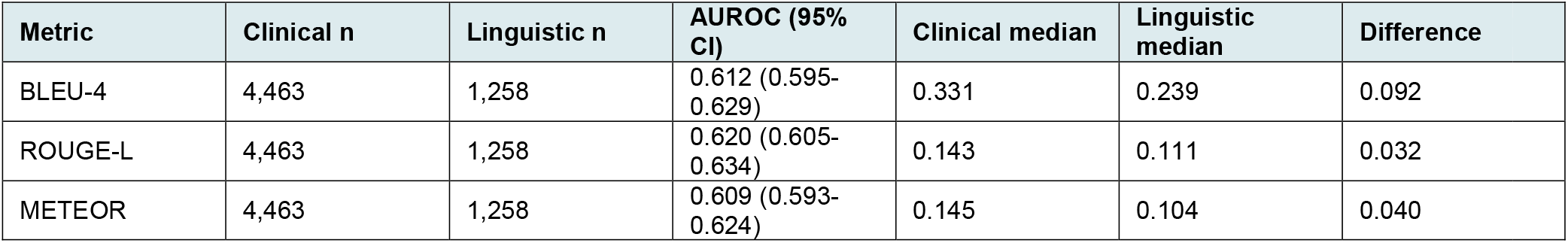
ReXErr-v1 clinical-content versus linguistic error selectivity.

**Figure 2.**
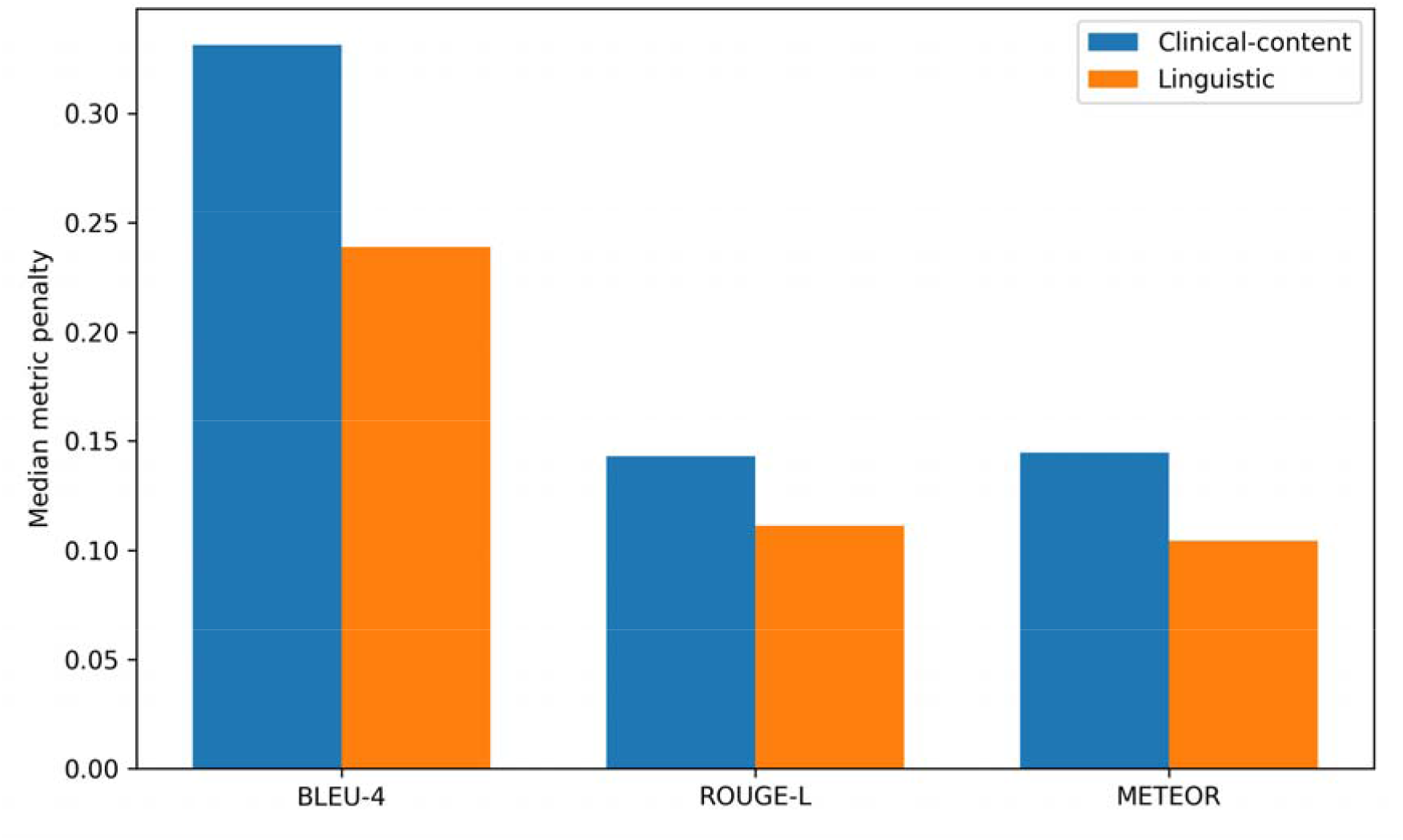
Median metric penalties for clinical-content and linguistic ReXErr-v1 errors. Clinical-content errors received larger median penalties than linguistic errors for BLEU-4, ROUGE-L and METEOR. Median penalties were 0.331 versus 0.239 for BLEU-4, 0.143 versus 0.111 for ROUGE-L, and 0.145 versus 0.104 for METEOR. Despite these differences, discrimination between clinical-content and linguistic errors remained modest. **Alt text:** Grouped bar chart comparing median penalties assigned by BLEU-4, ROUGE-L and METEOR to clinical-content and linguistic ReXErr-v1 errors; clinical-content penalties are higher for all three metrics.

In the clustered rank-penalty model, normalised character edit distance was strongly associated with penalty magnitude (coefficient 0.746; 95% CI 0.705-0.788; P<0.001). Original sentence length was negatively associated with penalty, whereas absolute token change was positively associated. Penalty magnitude was therefore strongly associated with the extent of textual change after adjustment for error type.

### Alignment with expert errors in RadEvalX

RadEvalX contained 151 clinically significant and 111 clinically insignificant errors. Omission of a finding accounted for 105 clinically significant and 66 clinically insignificant errors; several other categories were too sparse for reliable category-specific analysis.

CheXbert showed the strongest association with clinically significant error count (Spearman rho=0.413; 95% CI 0.223-0.578; Holm-adjusted P<0.001) (**Table 4; Figure 3**) and the highest discrimination for reports containing at least one clinically significant error (AUROC 0.742; 95% CI 0.638-0.836; AP 0.827) (**Table 4; Figure 4**). ROUGE-L was the strongest lexical metric (rho=0.269; 95% CI 0.062-0.457; adjusted P=0.040; AUROC 0.628). BERTScore, RadCliQ, RadGraph F1, BLEU-4 and METEOR did not show statistically significant correlations after Holm adjustment.

**Table 4.** RadEvalX alignment with expert clinically significant error assessment.

| Metric | Spearman rho (95% CI) | Holm P | AUROC (95% CI) | AP | rho insignificant |
| --- | --- | --- | --- | --- | --- |
| CheXbert | 0.413 (0.223-0.578) | <0.001 | 0.742 (0.638-0.836) | 0.827 | 0.103 |
| ROUGE-L | 0.269 (0.062-0.457) | 0.040 | 0.628 (0.514-0.739) | 0.716 | 0.119 |
| BERTScore | 0.195 (-0.007-0.386) | 0.257 | 0.607 (0.491-0.723) | 0.686 | 0.151 |
| RadCliQ | 0.188 (-0.019-0.380) | 0.257 | 0.595 (0.475-0.709) | 0.682 | 0.177 |
| RadGraph F1 | 0.159 (-0.044-0.352) | 0.342 | 0.574 (0.453-0.690) | 0.659 | 0.147 |
| BLEU-4 | 0.138 (-0.068-0.337) | 0.342 | 0.600 (0.482-0.713) | 0.690 | 0.085 |
| METEOR | 0.110 (-0.097-0.315) | 0.342 | 0.545 (0.427-0.661) | 0.657 | 0.181 |

AP, average precision; AUROC, area under the receiver operating characteristic curve; CI, confidence interval. Holm P values refer to the primary clinically significant-error correlation family.

**Figure 3.**
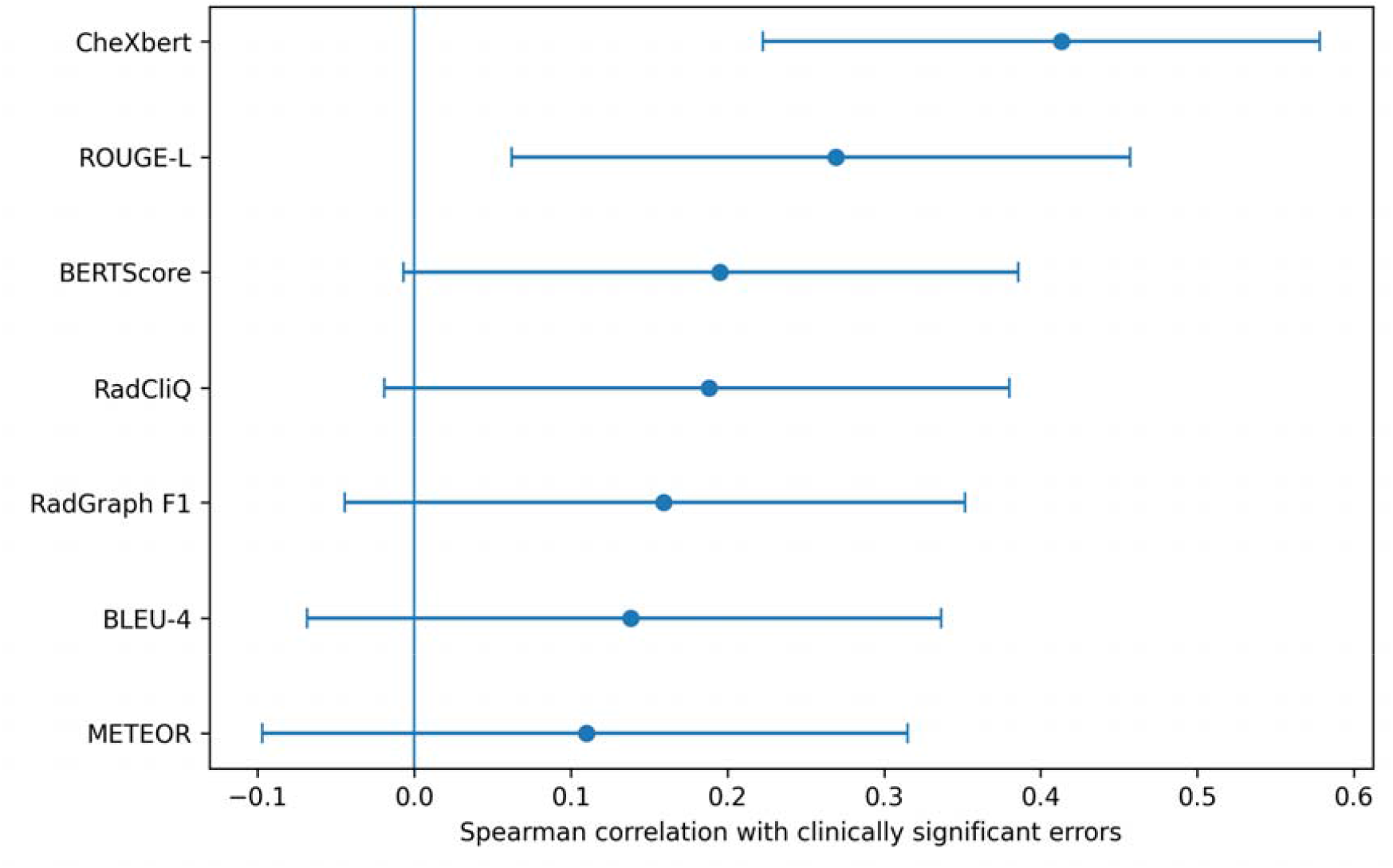
RadEvalX expert alignment. Spearman correlations between oriented metric error signals and clinically significant error counts. Points indicate correlations; horizontal bars indicate 95% bootstrap confidence intervals. **Alt text:** Forest plot comparing seven metrics’ Spearman correlations with clinically significant RadEvalX error counts; CheXbert is highest, while several confidence intervals cross zero.

**Figure 4.**
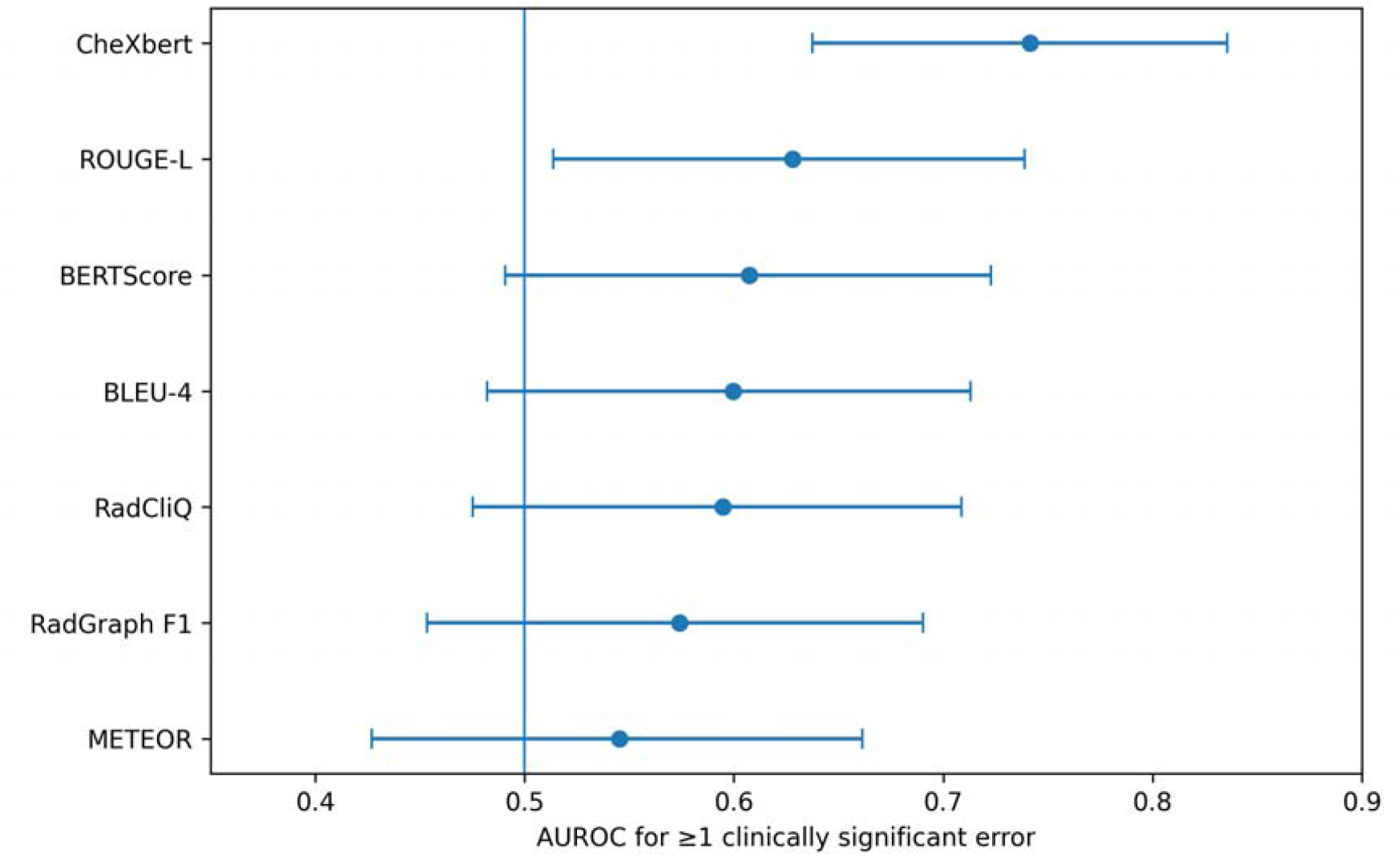
RadEvalX clinically significant error discrimination. AUROC values for detecting reports containing at least one clinically significant error. Points indicate AUROCs; horizontal bars indicate 95% bootstrap confidence intervals. **Alt text:** Forest plot comparing seven metrics’ AUROCs for detecting at least one clinically significant RadEvalX error; CheXbert is highest, while METEOR is lowest.

CheXbert was the only metric with a clearly positive difference between correlation with clinically significant and clinically insignificant errors (difference 0.311; 95% CI 0.003-0.604). In paired metric comparisons, CheXbert outperformed METEOR for correlation with clinically significant errors (difference 0.303; adjusted P=0.0126) and for AUROC (difference 0.196; adjusted P=0.0042); other pairwise differences did not remain significant after adjustment.

### Cross-dataset consistency

Among BLEU-4, ROUGE-L and METEOR, rank agreement between ReXErr-v1 sentence-level win rate and RadEvalX clinically significant-error correlation was only moderate (Spearman rank correlation 0.50; Kendall tau=0.33). With only three common metrics, these estimates are descriptive rather than inferential.

## Discussion

This cross-dataset analysis demonstrates an important distinction between alteration sensitivity and clinical selectivity. BLEU-4, ROUGE-L and METEOR detected almost every complete ReXErr-v1 modification, yet their ability to distinguish clinical-content from linguistic errors was weak. Normalised character edit distance was strongly associated with penalty magnitude, suggesting that a metric may appear highly sensitive because it responds to the extent of textual alteration, including spelling or homophone changes, without preferentially weighting clinically consequential discrepancies.^13,14^

The expert-annotated RadEvalX results reinforce this interpretation. CheXbert, a radiology-specific representation based on structured chest radiograph findings,^6^ showed the strongest available alignment with clinically significant error burden and the highest AUROC, although its correlation remained moderate and its confidence interval was wide. ROUGE-L was the strongest lexical comparator but showed lower expert alignment. These findings support concerns that conventional text-generation metrics can reward surface agreement without reliably measuring factual or clinical correctness, motivating radiology-aware and error-oriented evaluation strategies.^4,8-10,14^

RadEval has standardised access to lexical, semantic, clinical and large language model-based metrics and evaluated their correspondence with radiologist judgement. ReFINE and GEMA-Score have investigated interpretable and granular approaches to expert-aligned evaluation, with GEMA-Score reporting performance on RadEvalX.^11-12^ Particularly relevant, RadOT-Eval has evaluated both expert correlation on RadEvalX and corruption sensitivity on ReXErr-v1, demonstrating strong performance across these complementary settings.^13^

The present study therefore should not be interpreted as a competing metric leaderboard or as claiming novelty from combining RadEvalX and ReXErr-v1. Its contribution is instead to decompose metric behaviour into components not captured by aggregate benchmark performance. Alteration detection was separated from clinical selectivity; category-specific no-penalty analyses identified errors that metrics could fail to register; and the association between edit magnitude and penalty showed that apparent error sensitivity was strongly related to textual change. Together, these analyses show why strong corruption-detection performance or expert correlation does not establish preferential sensitivity to clinically consequential errors.^13,14^

The findings also highlight benchmark-specific limitations. ReXErr-v1 provides controlled perturbations at scale, but the present audit identified structural problems for sentence-level similarity analysis: 2,999 error-labelled rows lacked one side of a conventional pair, and 3.88% of nominal no-error rows contained non-identical text. The ReXErr-v1 custodians caution that sentence misalignment and incompletely generated errors occur and that synthetic errors should not be used as a sole benchmark.^15,16^ At report level, the fixed injection of three sampled errors prevents clean attribution of a report-level penalty to an individual error type. These characteristics support analysing perturbation behaviour rather than treating aggregate corruption discrimination as evidence of clinical validity.

For report-generation systems, these observations argue against reporting a single aggregate similarity score as evidence of clinical safety. A more defensible framework would combine lexical similarity for wording fidelity, clinically structured or entity-aware metrics for findings, and explicit error-oriented evaluation against expert judgement.^4,7-10^ Recent frameworks including RadEval, RadOT-Eval, ReFINE and GEMA-Score demonstrate progress toward broader and more clinically informed evaluation.^10-13^ The present findings suggest that such metrics should also be characterised by what drives their penalties, whether they distinguish clinically important from superficial perturbations, and which clinically relevant errors they fail to penalise.^13,14^

This study has several strengths. It combines a large synthetic perturbation benchmark with an independent expert-annotated dataset;^15-17^ uses paired report- and sentence-level analyses with parent-report bootstrap resampling; distinguishes clinically significant from clinically insignificant expert errors; and audits missing pairs, label noise, multiplicity and model non-convergence. The analysis also examines clinical-versus-linguistic selectivity, category-specific no-penalty failures and the relationship between textual edit magnitude and metric penalty. The local BLEU-4 implementation was verified against supplied RadEvalX values.

The study has limitations. First, it was not designed as a comprehensive benchmark of contemporary radiology report-evaluation metrics. ReXErr-v1 analyses were limited to BLEU-4, ROUGE-L and METEOR, while RadEvalX additionally included supplied BERTScore, CheXbert, RadGraph F1 and RadCliQ scores. RaTEScore and GREEN were not evaluated because their scores were unavailable in the supplied materials and reproducible implementations were not run within the analysis environment. The findings should therefore be interpreted as a failure-mode analysis rather than a ranking of contemporary metric performance. Second, the advanced RadEvalX scores were analysed as supplied rather than independently recomputed. Third, METEOR was calculated without WordNet synonym matching. Fourth, RadEvalX contains only 100 reports and several error categories were sparse, limiting category-specific inference. Fifth, sentence-level analyses were restricted to complete pairs, with missingness concentrated in particular error types; consequently, category-specific estimates may not represent the full ReXErr-v1 error distribution. Unpaired insertion/deletion errors and repetition were excluded from the clinical-versus-linguistic comparison. Finally, because the official test files were available before formal OSF registration, this work is exploratory rather than prospectively preregistered.

The practical implication is not that one metric should replace all others. Automated report evaluation should instead be treated as a measurement problem in which metrics capture different aspects of report quality and exhibit different failure modes. Near-ceiling synthetic corruption detection should not be equated with clinical validation, just as correlation with aggregate expert error burden does not establish sensitivity to every clinically important error category. Metrics intended for safety-sensitive model selection should therefore be evaluated by overall expert alignment, preferential penalisation of clinically consequential discrepancies, robustness to superficial linguistic perturbations and avoidance of clinically important no-penalty failures across independent datasets.

## Conclusion

Automated report-evaluation metrics were highly sensitive to textual corruption but differed substantially in alignment with clinical error importance. The available lexical metrics almost always detected ReXErr-v1 alterations yet only weakly separated clinical-content from linguistic errors, while CheXbert showed the strongest but still moderate alignment with clinically significant RadEvalX errors. Complementary, error-aware evaluation is therefore preferable to reliance on a single aggregate similarity metric.

## Data Availability

ReXErr-v1 version 1.0.0 is available through PhysioNet (doi:10.13026/9dns-vd94). RadEvalX version 1.0.0 is available through PhysioNet (doi:10.13026/tp88-q278). The present manuscript analyses only data contained in these released datasets and the supplied metric-score files.

