## Supplementary Material 1 for "Clinical selectivity and failure modes of automated chest radiograph report evaluation metrics: a cross-dataset analysis of ReXErr-v1 and RadEvalX"

### Supplementary Methods

This supplement provides the complete computational implementation underlying the statistical analyses reported in the manuscript. It expands technical details that were abbreviated in the main Methods section and does not introduce additional outcomes.

#### S1. Software environment and implementation

All analyses were performed in Python 3.13.5. No R code was used. A fixed NumPy random-number-generator seed of 20260806 was used for bootstrap resampling. Metric scores were retained at full floating-point precision throughout calculation, ranking, resampling and hypothesis testing; rounding was applied only for presentation in tables and text.

| **Component** | **Package** | **Version** | **Exact function(s)** | **Role** |
| --- | --- | --- | --- | --- |
| Runtime | Python | 3.13.5 | — | Analysis runtime |
| Arrays/random sampling | NumPy | 2.3.5 | default_rng(); integers(); quantile() | Random sampling and bootstrap percentiles |
| Data management | pandas | 2.2.3 | read_csv(); merge(); groupby(); Series.rank() | Import, linkage, grouping and within-metric ranks |
| Correlation | SciPy | 1.17.0 | scipy.stats.spearmanr() | Spearman rank correlations |
| Discrimination | scikit-learn | 1.8.0 | roc_auc_score(); average_precision_score() | AUROC and average precision |
| Regression | statsmodels | 0.14.6 | statsmodels.formula.api.gee(); Gaussian(); Exchangeable() | GEE modelling |
| Multiplicity | statsmodels | 0.14.6 | statsmodels.stats.multitest.multipletests(method='holm') | Holm adjustment |
| BLEU/METEOR | NLTK | 3.9.2 | sentence_bleu(); single_meteor_score(); PorterStemmer() | Lexical metric computation |
| ROUGE/edit distance | RapidFuzz | 3.14.3 | LCSseq.similarity(); Levenshtein.distance() | ROUGE-L longest common subsequence and character edit distance |
| Figures | Matplotlib | 3.10.8 | matplotlib.pyplot | Figure generation |

#### S2. Metric-score precision, penalty and numerical ties

For a higher-is-better metric, the paired penalty was defined as Δmi = Qoriginal,mi − Qerror,mi, where Qoriginal,mi is the score assigned to the original text and Qerror,mi is the score assigned to the error-injected text against the same reference. Scores were never rounded before calculation.

The numerical tie tolerance was ε = 1×10−12. Pairwise outcome Wmi was assigned as 1 when Δmi>ε, 0.5 when |Δmi|≤ε, and 0 when Δmi<−ε. The pairwise win rate was the arithmetic mean of Wmi across eligible pairs. The no-penalty indicator was I(Δmi≤ε), and therefore included both ties and any case in which an erroneous text received a higher quality score than its original counterpart.

#### S3. Lexical metric implementations

BLEU-4 was calculated with nltk.translate.bleu_score.sentence_bleu() using lower-cased whitespace-tokenised text, equal 1/4 weights for one- to four-grams and no smoothing. The locally recomputed RadEvalX BLEU-4 scores matched the supplied values to a maximum absolute difference below 5×10−10.

ROUGE-L used the longest common subsequence (LCS) length returned by rapidfuzz.distance.LCSseq.similarity(). For an original/reference token sequence of length r and candidate sequence of length c, recall=LCS/r, precision=LCS/c and ROUGE-L F1=2×precision×recall/(precision+recall), with zero assigned when the denominator was zero.

METEOR used nltk.translate.meteor_score.single_meteor_score() on lower-cased whitespace-tokenised text with exact matching and Porter stemming via nltk.stem.porter.PorterStemmer(). WordNet synonym matching was not used because the WordNet resource was unavailable in the analysis environment; the reported values therefore refer specifically to this exact-and-stem implementation.

#### S4. Bootstrap implementation and confidence intervals

All reported bootstrap confidence intervals used 10,000 non-parametric replicates generated with numpy.random.default_rng(20260806). ReXErr-v1 resampling was clustered at the parent-report level: for K unique parent reports, K report identifiers were sampled with replacement and all eligible sentence observations from each sampled report were carried together into the replicate. This preserved within-report dependence and prevented individual sentences from being resampled as independent observations.

RadEvalX resampling used the individual report/study as the bootstrap unit. Each replicate sampled 100 reports with replacement. Statistics were recomputed from each resampled dataset. Ninety-five per cent confidence intervals were percentile intervals defined by the 2.5th and 97.5th percentiles of the 10,000 bootstrap estimates, implemented with numpy.quantile(). Bias-corrected and accelerated intervals were not used.

#### S5. Within-metric rank-normalised penalty and GEE

Because raw penalties have different scales across BLEU-4, ROUGE-L and METEOR, the continuous GEE outcome was constructed by ranking raw penalty values separately within each metric using pandas.Series.rank(method='average', pct=True). Thus, for metric m with Nm observations, Rmi=average_rank(Δmi)/Nm. Tied penalties received their average rank. This was a percentile-rank transformation, not an inverse-normal transformation, z-score or min-max scaling.

The final clustered model was a Gaussian GEE fitted with statsmodels.formula.api.gee(), Gaussian family and Exchangeable() working correlation structure. Parent report (study_id) was the clustering variable. The model included fixed effects for metric, error type, metric×error-type interaction, original sentence token count, absolute token change and normalised character edit distance. The default robust/sandwich GEE covariance estimator was used. BLEU-4 and 'Add typo' were the reference categories generated by categorical coding.

A binomial/logistic GEE for binary error detection was attempted but did not converge because ROUGE-L and METEOR detected nearly all eligible errors, creating quasi-complete separation. The Gaussian GEE of rank-normalised penalty was therefore retained as the continuous clustered analysis.

#### S6. Text-derived edit covariates

Tokenisation for the edit covariates was lower-cased whitespace splitting: tokens(s)=str(s).lower().split(). Original sentence length was the number of tokens in the original sentence.

Absolute token change was defined as |Nerror−Noriginal|, where Nerror and Noriginal are the lower-cased whitespace-token counts of the erroneous and original sentences, respectively. It was therefore an absolute count difference rather than a percentage or proportional change.

Normalised character edit distance was defined as Dchar=L(lower(original), lower(error))/max(|lower(original)|, |lower(error)|, 1), where L is the character-level Levenshtein distance calculated by rapidfuzz.distance.Levenshtein.distance(). A value of 0 indicates identical lower-cased character strings; larger values indicate more extensive character modification.

#### S7. RadEvalX expert-alignment analyses

The binary discrimination outcome was Yi=1 when the consensus clinically significant error count was >0 and Yi=0 when it was 0. For metrics in which higher values represented better report quality (BLEU-4, ROUGE-L, METEOR, BERTScore, CheXbert and RadGraph F1), the expert-alignment error signal was −Q. RadCliQ was retained in its supplied error-oriented direction.

AUROC was calculated with sklearn.metrics.roc_auc_score(Y, error_signal). Precision-recall performance was calculated with sklearn.metrics.average_precision_score(Y, error_signal). This quantity is average precision (AP), not trapezoidal integration of the precision-recall curve; the manuscript therefore reports it as AP rather than AUPRC.

Spearman correlations were calculated with scipy.stats.spearmanr(). For each metric, ρsig correlated the oriented error signal with clinically significant error count, and ρinsig correlated it with clinically insignificant error count. The selectivity contrast was Δρ=ρsig−ρinsig.

#### S8. Bootstrap contrasts and paired metric comparisons

Correlation differences were bootstrapped in a paired manner. Within each RadEvalX bootstrap replicate, the same resampled reports were used to calculate both ρsig and ρinsig, after which Δρ(b)=ρsig(b)−ρinsig(b) was recorded. Its 95% confidence interval was the percentile interval of the 10,000 paired differences.

For pairwise comparison of metrics A and B, a single set of bootstrap report indices was generated for each replicate and applied to both metrics. Correlation and AUROC contrasts were Dρ(b)=ρA(b)−ρB(b) and DAUC(b)=AUROCA(b)−AUROCB(b), respectively. Two-sided empirical bootstrap P values were calculated as 2×min[Pr(D(b)≤0), Pr(D(b)≥0)], capped at 1.0. Pairwise P values were Holm-adjusted separately for the correlation and AUROC comparison families using statsmodels.stats.multitest.multipletests(method='holm').

#### S9. Missing-data and linkage rules

No statistical imputation was performed. ReXErr-v1 report-level analyses required both original_report and error_report. The primary sentence-level error analysis required error_present=1 and non-missing original_sentence and error_sentence. Of 8,723 error-labelled sentence rows, 2,523 lacked an original sentence and 476 lacked an altered sentence, leaving 5,724 complete pairs. Missing text was never converted to the literal string 'nan' or an empty string for metric calculation.

The clinical-versus-linguistic grouped analysis used 4,463 clinical-content and 1,258 linguistic pairs. Three complete add-medical-device pairs were retained in overall and category-specific analyses but excluded from the grouped comparison because the category was too sparse for a stable grouped estimate.

The RadEvalX clinically significant and clinically insignificant error files were merged by report_id. Blank error-category cells were interpreted as zero errors and were filled with zero before category counts were summed. The supplied metric-score file was linked by report_id; all 100 expert-reviewed reports matched. A metric-specific missing score, if present, was excluded only from analyses involving that metric, and a paired comparison between two metrics required availability of both metric scores on the same report.

#### S10. Reproducibility notes

The analyses were not prospectively preregistered and are therefore considered exploratory secondary analyses rather than prospectively preregistered confirmatory analyses. BERTScore, CheXbert, RadGraph F1 and RadCliQ were analysed as supplied and were not independently recomputed. RaTEScore and GREEN were not evaluated because their scores were unavailable in the supplied materials and reproducible implementations were not run within the present analysis environment. The study therefore does not represent a comprehensive benchmark of contemporary report-evaluation metrics.
